# Non communicable diseases and resistant tuberculosis, a growing burden among people living with HIV in Eastern Kenya

**DOI:** 10.1101/2025.01.08.25320218

**Authors:** Patrick Kiogora Muriuki, Musa Otieno Ngayo, Moses Njire, Juster Mungiria, Winnie Nyanya, Daniel Owuor, Perpetual Ndung’u

## Abstract

Human Immunodeficiency Virus (HIV) and tuberculosis (TB) still poses a significant health burden in Kenya. Countries with the highest burden of people living with HIV (PLWH) have a substantial burden of non-communicable diseases (NCDs) including type 2 diabetes (T2D) and hypertension (HPT). This study evaluated the burden and predictors of T2D, HPT and TB including resistant strains among PLWH receiving antiretroviral therapy (ART) in Eastern Kenya. Blood and sputum samples were obtained from 280 consenting PLWH and a detailed sociodemographic questionnaire was administered. TB and rifampicin resistance was determined by Cepheid’s GeneXpert system while Glycated hemoglobin (HbA1c) was determined using SD A1cCare™ analyzer. Blood pressure (BP) measurements with systolic BP readings of≥140 mmHg and/or diastolic BP≥90 mmHg was considered hypertensive. The participants CD4 cell counts, plasma HIV-1 RNA, full blood cells and blood chemistry were also measured. The patients mean (SD) age was 35.6 (±9.8) years, with 169 (82.4%) being on the first line ART regimen and a median (IQR) duration living with of 7 (4 to 8) years. The majority of the patients 179 (63.9%) were HIV mono-infected. The dual coinfections reported included; 58 (20.7%) HIV/TB, 42 (15%) HIV/ T2D, 33 (11.8%) HIV/HPT. Triple coinfections included; 18 (6.4%) HIV/T2D/HPT, 9(3.2%) HIV/TB/T2D, 9 (3.2%) HIV/TB/HPT with quadruple coinfection of HIV/TB /T2 /HPT among 4 (1.4%) patients. Six 6 (2.1%) HIV patients were coinfected with multidrug resistant TB. In the multivariable model, being on ARV only (aOR 0.5; 95% CI 0.4 – 0.6, p = 0.0001) and virological suppression (aOR 0.8; 95% CI 0.6 – 0.9, p = 0.017) were protective to HIV/TB coinfection. Previously hospital admission (aOR 1.2; 95% CI 1.1 – 1.4, p = 0.049) and previous TB infection (aOR 1.6; 95%CI 1.0 – 3.0, p = 0.034) were associated with HIV/TB coinfection. This study shows that Eastern Kenya is experiencing a syndemic of NCDs and TB including resistant strains among PLWH denoting the need to integrate the management of NCDs among HIV and TB treatment programs in Kenya.

## Introduction

Human Immunodeficiency Virus (HIV) is still a public health threat in Kenya, by 2023 about 1.4 million people were living with HIV (PLWH) [1]. Since the initiation of the antiretroviral therapy (ART) in 1990s and the subsequent scale-up, HIV in Kenya is currently a manageable chronic condition. About 94% of PLWH are currently on ART with 94% achieving virological suppression. Increased life expectancy among PLWH has become associated with the concurrent increase in the prevalence of both infectious diseases such as Tuberculosis (TB) as well as non-communicable diseases (NCDs) such as obesity, hypertension (HPT) and type 2 diabetes (T2D) [2]. There is a co-existence of both chronic infectious and non-infectious diseases, both exacerbated by a highly unequal society, poverty and other social determinants, and which reflect an epidemiological transition [3]. Tuberculosis is a major cause of ill health and death globally, prior to coronavirus (coViD-19) pandemic, TB ranked highest leading cause of mortality from a single infectious agent, ranking above HIV/AIDS [4]. By 2023 Kenya was among the African countries in the three global lists of high-burden countries for TB, HIV-associated TB and multidrug-resistant or rifampicin-resistant TB (MDR/RR-TB) [4]. In Kenya, the interaction between TB and HIV infection has increased the burden of TB with a prevalence of 237 per 100,000 population in 2022 and new estimated cases of 288 per 100,000 population in the same year. Over 7,000 and 8900 deaths among HIV negative and HIV positive respectively were attributed to TB in the same year [4].

In Sub-Saharan Africa, where the majority of NCDs have long been considered “diseases of affluence”, T2D and HPT are becoming increasingly prevalent [5, 6]. In Kenya, NCDs are estimated to account for 39% of all deaths, with HPT and T2D accounting for 13.8% and 3.5% of these deaths respectively [7, 8]. With improved socioeconomic status, reduced morbidity and mortality due to infectious diseases, rapid urbanization, and changes in lifestyle and dietary habits, developing countries are rapidly experiencing an increase in NCDs [9]. The TB infection only takes the form of disease as the immunity system of a person gets compromised. The NCDs are shown compromise the immunity of affected person. Prolonged dependence on medication, diabetes, smoking, and consumption of alcohol includes the major causes of TB [10]. Chronic TB infection have been shown to contribute to the development of HPT and vise verse, a reverse association may possibly exist, such that HPT may lead to an increased risk of developing TB [11]

Comorbidity of TB with NCDs such as T2D, chronic respiratory diseases, cardiovascular diseases, and cancers and other communicable diseases such as HIV/AIDS is prevalent in regions of the world that are highly endemic for tuberculosis and thus integrated and efficient responses are required to tackle these comorbidities in resource-poor countries [4]. There is no hope for an end to TB without tackling NCDs as a matter of urgency [12]. Even though the rate of active TB is much higher in specific groups, such as those on immunosuppressants or with HIV infection, as well as patients with comorbidities like T2D or HPT [13]. The risk of progression from latent infection to active disease in the general population is affected by factors such as cigarette smoking, alcohol consumption, low body mass index, physical inactivity, and malnutrition [10]. It is imperative that studies continue to provide data on the comorbidity of TB and lifestyle disease especially in developing countries in-order to strategize on the mean of reduce the burden of these co-infections. Therefore, understanding the associations of combined lifestyle risk factors with tuberculosis risk may be more informative for translating epidemiological findings into prevention strategies. This study therefore determined the burden and comorbidity of tuberculosis and lifestyle diseases among PLWH, Meru County Eastern Kenya.

## Methods

### Study design

This cross-sectional study was conducted among PLWH between 10^th^ June 2021 and 9^th^ June 2022 in Nyambene Sub-County Hospital in Meru County in Eastern Kenya

#### Eligibility criteria

Patients were recruited in this study if they were: (i) aged above 18 year (ii) consenting to the study, (iii) willing to provide sputum and blood samples (iv) willing to undergo clinical medical examination, and (v) had been receiving ART treatment and care at Nyambene Sub-County Hospital in Meru County in Eastern Kenya

#### Informed consent

Written informed consent was obtained from all subjects before the study.

#### Sample size

Sample size calculation used the formula described by Lemashow [14] based on population proportion estimation. Setting the alpha (α) at 0.05, critical value (z) based on the desired confidence level (∝ at 1.96), and the prevalence (p) of TB and lifestyle diseases in PLWH prevalence of 19.8% [15], a total of 280 eligible PLWH were recruited with 15% added to cover for lost to follow up.

#### Ethical statement

This research was carried out in accordance with the basic principles defined in the Guidance for Good Clinical Practice and the Principles enunciated in the Declaration of Helsinki (Edinburg, October 2000). This protocol and all the tools including informed consent forms were reviewed, and permission obtained from the Jomo Kenyatta University of Agriculture and Technology (JKUAT) Ethical Review Committee (ERC) JKU/IERC/02316/0134 while the study license obtained from the National Commission For Science,Technology & Innovation (NACOSTI) (License No. NACOSTI/21/12006). Written informed consent was obtained from all patients before enrollment.

#### Data collection

A detailed, structured, face-to-face interview gathered information on patient’s socio-demographic, ARV use and medical history. Blood samples (5 mL) were collected into two blood tube as follows: EDTA anticoagulant tube for immunological testing as well as hematological analysis while serum separating tube for clinical chemistry. The samples were stored at −80 C° after collection awaiting analysis.

Early morning two sputum samples were collected from each patient. For patients who were unable to expectorate the sputum, ultrasonic nebulizer technique was used for sputum induction. The samples were held at 4^°^C until processed by standard laboratory procedures and the Xpert assay.

The patients’ blood pressure (BP) was measured using Citizen Digital Arm Blood Pressure Monitor CH-503 (Citizen Systems Japan CO., LTD). Blood pressure (BP) measurements with systolic BP readings of≥140 mmHg and/or diastolic BP≥90 mmHg was considered hypertensive.

### Laboratory analysis

#### Glycated haemoglobin (HbA1C) Testing

Glycated haemoglobin (HbA1C) testing was used to determine the mean blood sugar concentration and the degree of carbohydrate imbalance over the preceding two to three months. The HbA1C was measured using SD A1cCare™ analyzer (SD Biosensor, Inc). The test interpretation was as follows; <8% HbA1c (64mmol/mol) Less stringent glycemic control, < 7% HbA1c (53mmol/mol) General glycemic control, while < 6.5% HbA1c (48mmol/mol) means more stringent glycemic control. As recommended by the ADA, patients in the range of 5.7%-6.4% HbA1c (39-46 mmol/mol) would be in the category of increased risk for diabetes.

### Detection of Tuberculosis

#### Acid- Fast Bacillus (AFB) smear microscopy

Ziehl-Neelsen staining was performed on the first non-decontaminated sputum samples. Liquid samples were first concentrated for 15min at 3000rpm and sediments were used. Purulent sputum was liquefied with N acetyl-L-cysteine as a mucolytic agent to increase the homogeneity of the sample before smear preparation. Smears were examined to explore the presence of acid-fast bacilli and graded as per the International Union against Tuberculosis and Lung Disease scale; negative for TB, scanty, + 1, +2, and + 3. A patient was considered positive if a minimum of one smear was graded scanty or higher.

#### Detection of MTB/RIF

The second unprocessed sputum were collected in specialized containers and tested directly using the GeneXpert MTB/RIF diagnostic system (Cepheid, Sunnyvale, CA, USA) as described by Fouda et al., [16]. One ml un-concentrated specimens (without centrifuge) were used for Xpert MTB/RIF assay. Sample reagent (two volumes of 0.1M NaOH and 0.1M isopropanol) was added in a 2:1 ratio to unprocessed specimen in falcon tube and the tube was manually agitated twice during a 15min incubation period at room temperature. Subsequently, 2ml of the inactivated sample was transferred to the labeled test cartridge by a sterile disposable pipette and loaded into The Xpert MTB/RIF instrument. The assay uses nucleic acid probes that identify and report the presence or absence of the normal, rifampicin-susceptible, sequence of the *rpoB* gene of MTB. Five different colored beacons are used, each covering a separate nucleic acid sequence within the amplified *rpoB* gene. The results of the assay are: a-TB positive rifampicin resistant, b-TB positive rifampicin non resistant, c-TB not detected and d-Invalid result [16].

### Immunological and Biochemical assessment

The CD4 counts was measured as absolute numbers and percentages using a BD FACSpresto^TM^ (Becton Dickinson, BD Biosciences, San Jose, USA). Full hemogram was measured by the Medonic M - series differential hematology analyzer (Boule Clinical Diagnostic Solutions, Inc.). Blood clinical chemistry (AST, ALT, Creatinine) was measured using the DRI-CHEM NX500 dry chemistry analyzer (Fujifilm, Czech Republic). All these assays were done according to the manufacturer’s instruction and the existing standard operating procedures.

### Determination of plasma HIV-1 RNA concentration

To measure the HIV-1 RNA concentration (viral load) involved, first the extraction of viral RNA from 1ml plasma samples of study patients using QIAmp viral RNA mini kit (Cat. No. 52906, Qiagen Inc., USA) in line with the manufacturer guidelines. A total volume of 10μL of HIV RNA was quantified using the Generic HIV Viral Load assay (Biocentric, Bandol-France). The amplification of HIV RNA was achieved using the ABI Prism 7300 Sequence Detection System (Applied Biosystems). The amplification condition was set at 50°C for 10 minutes and 95°C for 5 minutes, followed by 50 cycles of 95°C for 15 seconds and 60°C for 1 minute. The test has detection limit of 40 HIV-1 RNA copies/ml.

### Data analysis

Descriptive statistics were used to summarize the sociodemographic, socioeconomic, and clinical data. The proportion of participants with different comorbidity of TB and lifestyle diseases were calculated. Bivariable logistic regression analyses were conducted to assess the association of each variable (demographic and clinical) with NCDs, TB among PLWH Subsequently, a multivariable logistic regression model was used to explore the independent associations between the identified correlates and various comorbidity of TB and lifestyle diseases listed above. Data analysis was performed using R^®^ software version 4.1.2 at a significance level of p ≤0.05.

## Results

### Baseline Characteristics of Study Patients

Table 1 summarizes the baseline characteristics of the study population. Of the 280 PLWH recruited from Nyambene region, Meru County, their mean (SD) age was 35.6 (±9.8) years, 146 (52.1%) were male, 91 (32.5%) had no formal education level while 207(73.9%) were married. The majority of the participants 169 (82.4%) were on the first line ART regimen combination of Dolutegravir/lamivudine/tenofovir (DTG/3TC/TDF) with 179(63.9%) on while 4(1.4%) patients were on four different drug combinations (ARV + Anti-TB + antidiabetic + antihypertensive). The median (IQR) duration living with HIV of 7 (4 to 8) years and median (IQR) duration on ART of 6 (4 to 7). There were 28 (10%) of the participants who were hospitalised based on the current conditions with 104 (37.1%) non adherence rate. About half 143 (51.1%) of the participate had cough lasting > 3 weeks with 6 (2.1%), 22 (7.9%) and 22 (7.9%) previously diagnosed with tuberculosis, diabetes and hypertension respectively. There were 114 (50.7%) and 138 (50.7%) of the participants with family history of hypertension and diabetes respectively. Of the 280 recruited participants, 97 (34.6%) were taking alcohol while 78 (27.9%) were either smoking cigarettes or taking tobacco products.

**Table 1.**
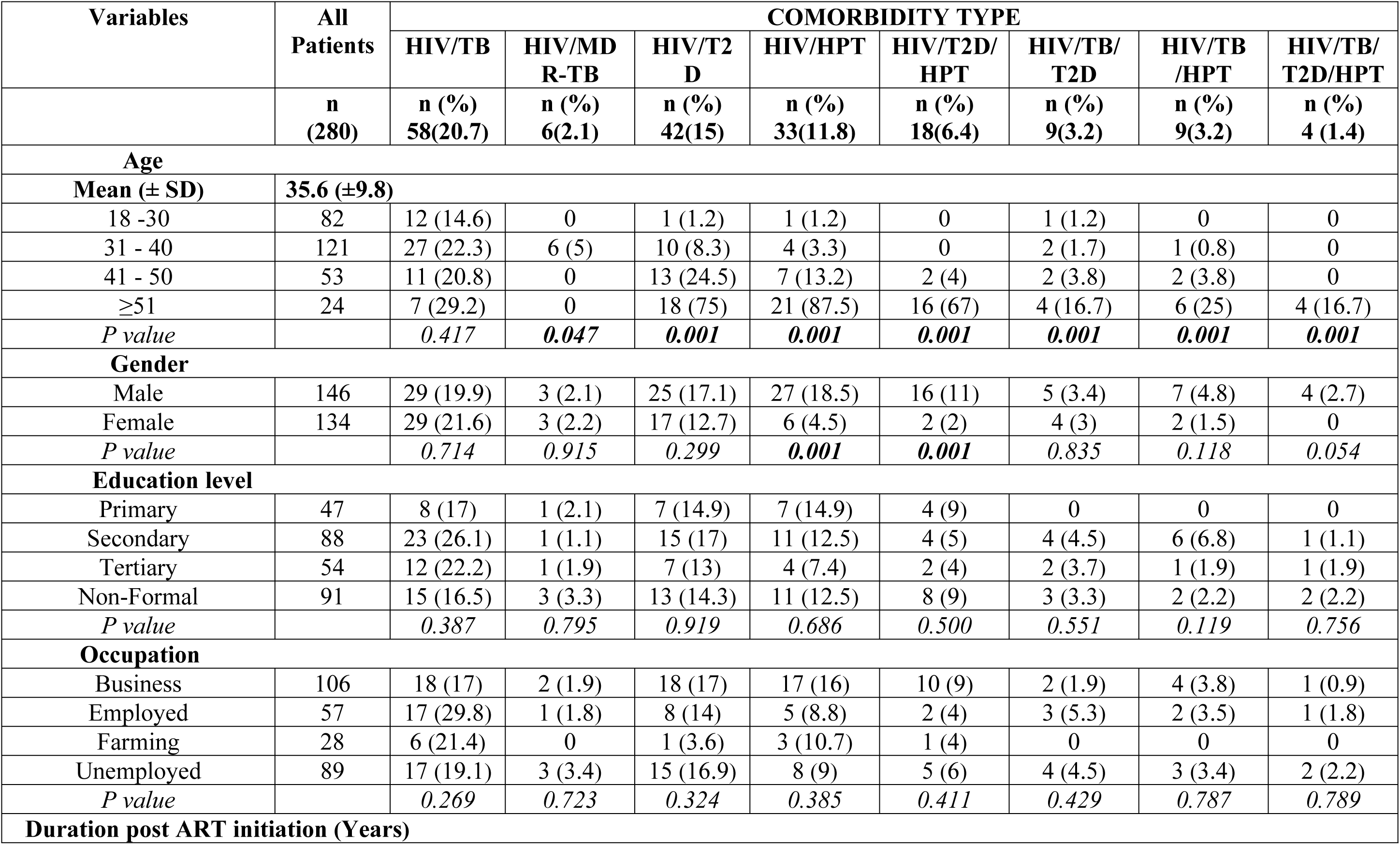
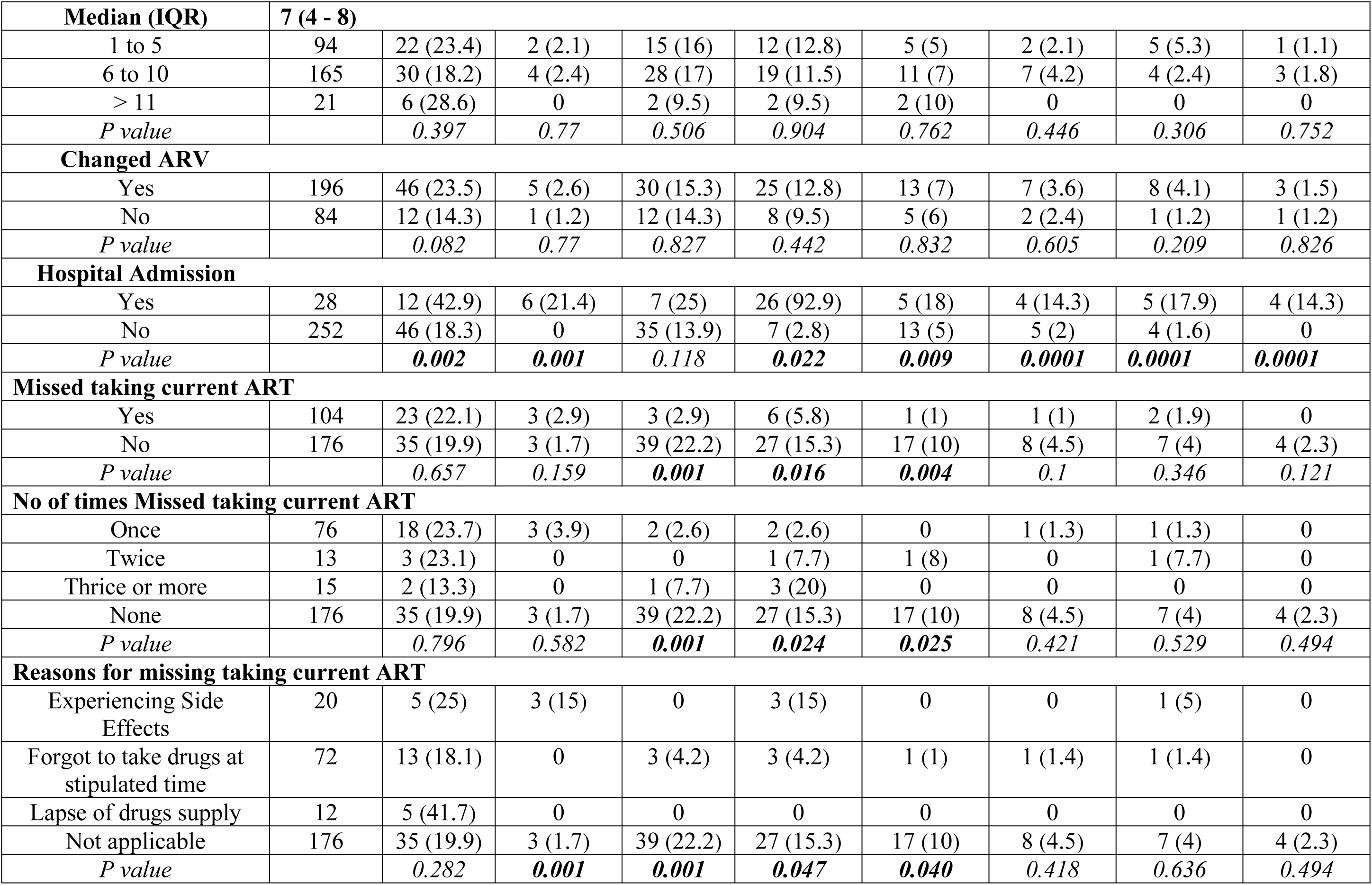
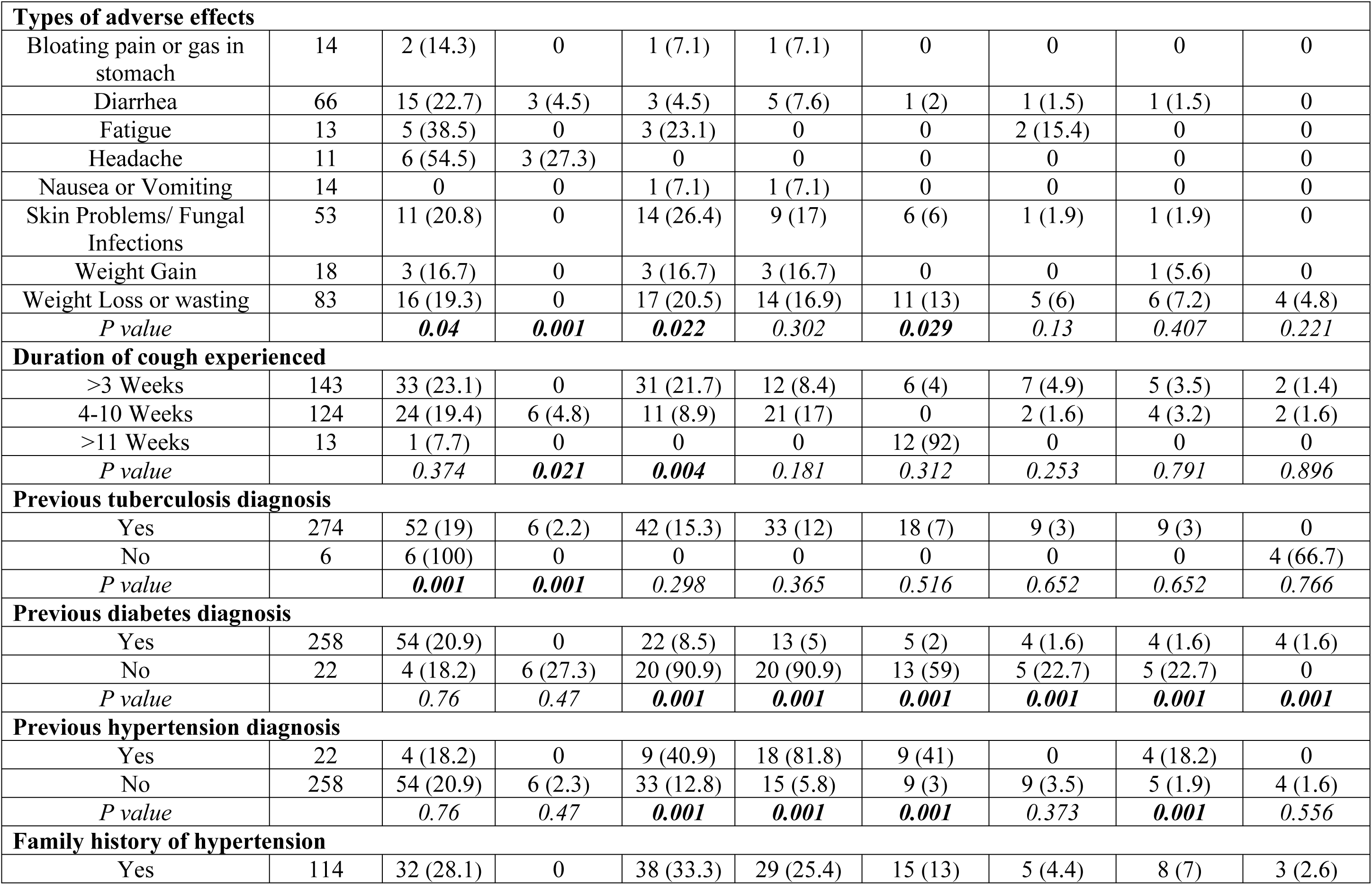
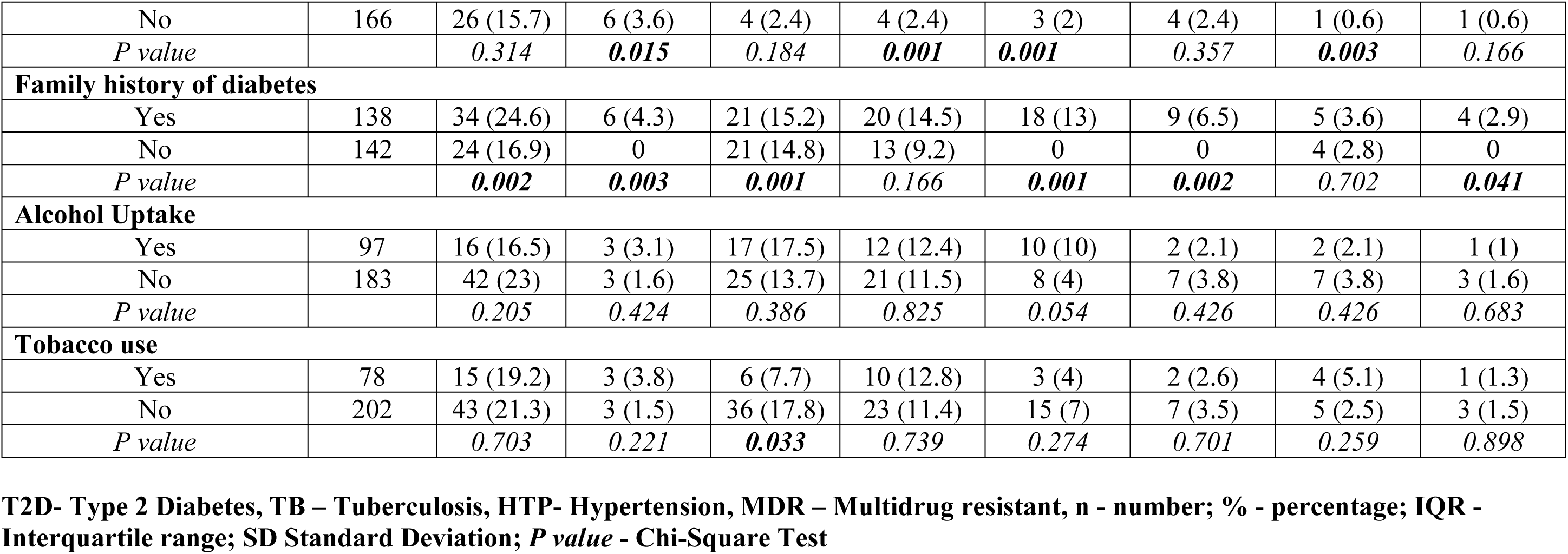
Baseline characteristics of the study population (n =280 patients)

### Immuno-pathological laboratory outcomes

Table 2 summarizes the immune-pathological characteristics of study participants. The mean (SD) BMI of the study participants was 25.7 (± 4.8) Kg/M^2^with the majority of them 132 (47.1) being within the normal weight category. There were 4 (1.4%) and 52 (18.6%) of the participants who were underweight and obese respectively. Majority of the participants 186 (66.4%) of the participants were immunologically responding to treatment ≥ 500 cells/mm^3^ while 246 (87.9%) were currently virally suppressed < 50 copies/mL. There were 26 (9.3%) and 52 (18.6%) with elevated liver enzymes (ALT and AST) respectively. About 83 (29.4%) had HB <13 (g/dL) considered anaemic while 141 (50.4%) had creatinine levels >0.8mg/dL considered elevated (Table 2)

**Table 2.** Summary of patient Immuno-pathological laboratory outcomes.

| Variables | All Patients | COMORBIDITY TYPE |  |  |  |  |  |  |  |
| --- | --- | --- | --- | --- | --- | --- | --- | --- | --- |
|  |  | HIV/TB | HIV/MDR-TB | HIV/T2D | HIV/HPT | HIV/T2D/HPT | HIV/TB/T2D | HIV/TB/HPT | HIV/TB/T2D/HPT |
|  | n (280) | n (%) 58(20.7) | n (%) 6(2.1) | n (%) 42(15) | n (%) 33(11.8) | n (%) 18(6.4) | n (%) 9(3.2) | n (%) 9(3.2) | n (%) 4 (1.4) |
| **Body mass index (Kg/m<sup>2</sup>)** |  |  |  |  |  |  |  |  |  |
| *Mean (± SD)* | **25.7 (± 4.8)** |  |  |  |  |  |  |  |  |
| <18.5 (Underweight) | 4 | 0 | 0 | 1 (25) | 0 | 0 | 0 | 0 | 0 |
| 18.5 to 24.9 (Normal weight) | 132 | 25 (18.9) | 4 (3) | 21 (16) | 66 (50) | 10 (8) | 5 (3.8) | 3 (2.3) | 2 (1.5) |
| 25 to 29.9 (Overweight) | 92 | 16 (17.4) | 1 (1.1) | 16 (17) | 17 (18) | 1 (1) | 2 (2.2) | 2 (2.2) | 1 (1.1) |
| ≥30 (Obese) | 52 | 17 (32.7) | 1 (1.9) | 4 (8) | 10 (19) | 7 (13) | 2 (3.8) | 4 (7.7) | 1 (1.9) |
| *P value* |  | 0.091 | 0.781 | 0.393 | 0.858 | 0.470 | 0.883 | 0.243 | 0.972 |
| **Baseline CD4+ (cell/μL)** |  |  |  |  |  |  |  |  |  |
| **Median (IQR)** | **424.5 (301.5 - 531.5)** |  |  |  |  |  |  |  |  |
| 1-500 | 186 | 38 (20.4) | 6 (3.2) | 29 (16) | 24 (13) | 14 (8) | 5 (2.7) | 4 (2.2) | 2 (1.1) |
| >501 | 94 | 20 (21.3) | 0 | 13 (14) | 9 (10) | 4 (4) | 4 (4.3) | 5 (5.3) | 2 (2.1) |
| *P value* |  | 0.869 | 0.078 | 0.697 | 0.415 | 0.292 | 0.483 | 0.156 | 0.483 |
| **HIV viral load (Cells/mls)** |  |  |  |  |  |  |  |  |  |
| *Mean (± SD)* | **6404.8 (± 29077.7)** |  |  |  |  |  |  |  |  |
| 1-1000 | 246 | 44 (17.9) | 0 | 41 (17) | 32 (13) | 18 (7) | 9 (3.7) | 8 (3.3) | 4 (1.6) |
| >1001 | 34 | 14 (41.2) | 6 (17.6) | 1 (3) | 1 (3) | 0 | 0 | 1 (2.9) | 0 |
| *P value* |  | **0.002** | **0.001** | **0.036** | 0.088 | 0.103 | 0.257 | 0.923 | 0.454 |
| **Baseline ALT (U/L)** |  |  |  |  |  |  |  |  |  |
| **Median (IQR)** | **24 (19 - 34)** |  |  |  |  |  |  |  |  |
| <56 | 254 | 54 (21.3) | 6 (2.4) | 38 (15) | 28 (11) | 16 (6) | 9 (3.5) | 8 (3.1) | 4 (1.6) |
| ≥56 | 26 | 4 (15.4) | 0 | 4 (15) | 5 (19) | 2 (8) | 0 | 1 (3.8) | 0 |
| *P value* |  | 0.481 | 0.428 | 0.954 | 0.216 | 0.783 | 0.329 | 0.923 | 0.454 |
| **Baseline AST (U/L)** |  |  |  |  |  |  |  |  |  |
| **Median (IQR)** | **25.3 (20.5 - 38)** |  |  |  |  |  |  |  |  |
| <40 | 228 | 49 (21.5) | 3 (1.3) | 34 (15) | 25 (11) | 12 (5) | 6 (2.6) | 6 (2.6) | 1 (0.4) |
| ≥40 | 52 | 9 (17.3) | 3 (5.8) | 8 (15) | 8 (15) | 6 (12) | 3 (5.8) | 3 (5.8) | 3 (5.8) |
| *P value* |  | *0.502* | ***0.045*** | *0.931* | *0.372* | *0.095* | *0.247* | *0.247* | ***0.003*** |
| **Baseline HB (g/dL)** |  |  |  |  |  |  |  |  |  |
| **Median (IQR)** | **14.3 (12.8 - 15.5)** |  |  |  |  |  |  |  |  |
| <13 | 83 | 16 (19.3) | 1 (1.2) | 9 (11) | 8 (10) | 4 (5) | 3 (3.6) | 3 (3.6) | 0 |
| ≥13 | 197 | 42 (21.3) | 5 (2.5) | 33 (17) | 25 (13) | 14 (7) | 6 (3) | 6 (3) | 4 (2) |
| *P value* |  | *0.7* | *0.482* | *0.206* | *0.47* | *0.476* | *0.805* | *0.805* | *0.191* |
| **Creatinine** |  |  |  |  |  |  |  |  |  |
| **Median (IQR)** | **0.9 (0.6 - 1.1)** |  |  |  |  |  |  |  |  |
| <0.8mg/dL | 139 | 26 (18.7) | 1 (0.7) | 24 (17) | 17 (12) | 8 (6) | 4 (2.9) | 4 (2.9) | 1 (0.7) |
| >0.8mg/dL | 141 | 32 (22.7) | 5 (3.5) | 18 (13) | 16 (11) | 10 (7) | 5 (3.5) | 5 (3.5) | 3 (2.1) |
| *P value* |  | *0.41* | *0.095* | *0.292* | *0.819* | *0.604* | *0.751* | *0.751* | *0.321* |

**T2D- Type 2 Diabetes, TB – Tuberculosis, HTP- Hypertension, MDR – Multidrug resistant, n - number; % - percentage; IQR -**

### Prevalence of TB and NCDs among PLWH

The majority of the patients 179 (n = 280; 63.9%) were HIV mono-infected. The following comorbidities were determine; 58 (n = 280; 20.7%) HIV/TB coinfection, 42 (n = 280; 15%) HIV / T2D, 33 (n = 280; 11.8%) HIV / HPT, 18 (n = 280; 6.4%) HIV / T2D/ HPT, 9 (n = 280; 3.2%) HIV / TB/ T2D, 9 (n = 280; 3.2%) HIV / TB/ HPT and 4 (n = 280; 1.4%) HIV / TB / T2D / HPT. There were 6 (n = 280; 2.1%) PLWH coinfected with multidrug resistant TB (MDR-TB). There was however no PLWH with multidrug resistant TB coinfected with either diabetes or hypertension (Fig 1).

**Figure 1.**
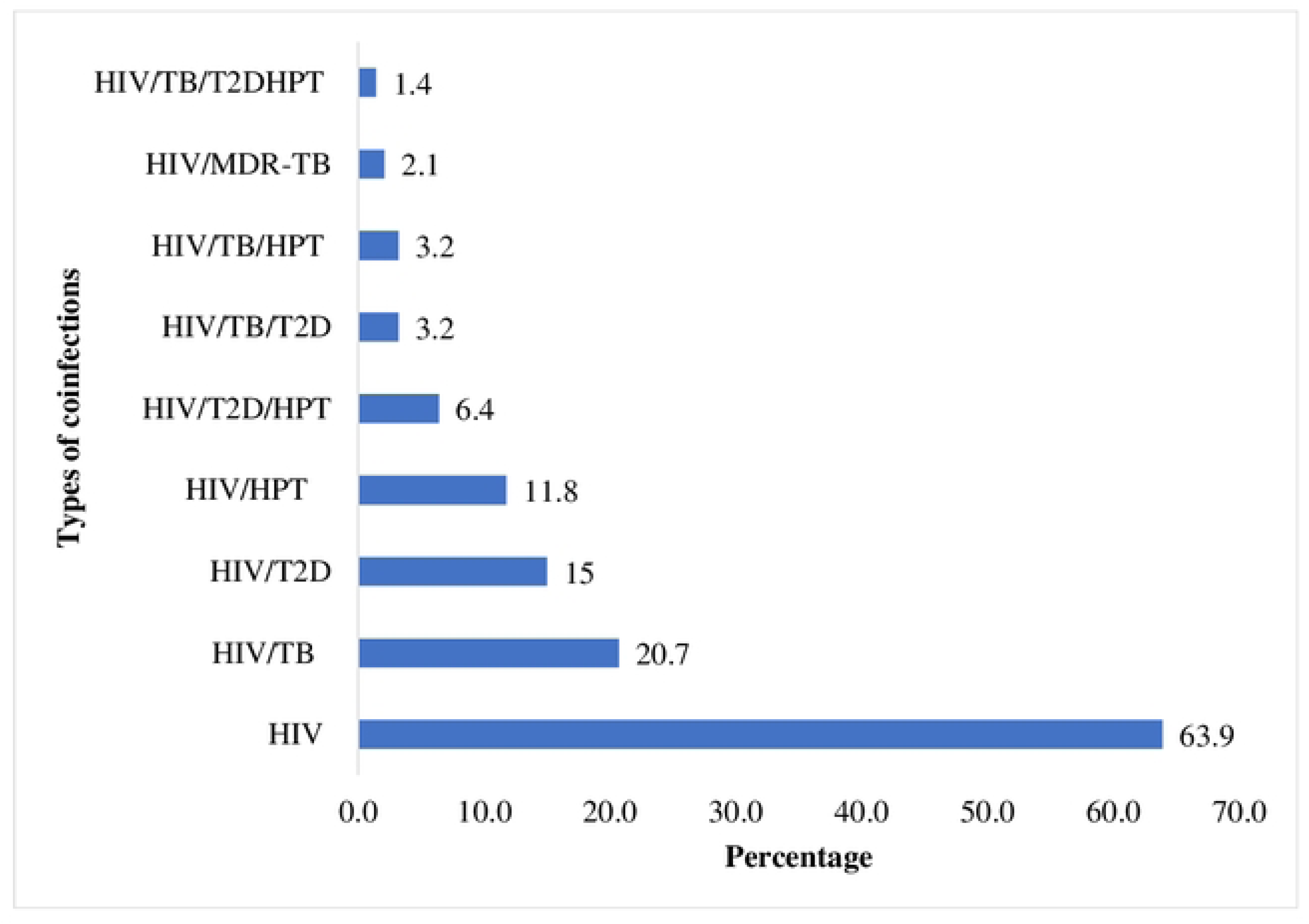
The distributions in the types of comorbidities among study participants

### Comparison Between Baseline Characteristics and HIV comorbidities

Univariate analysis of baseline characteristics revealed that participants age, previous hospital admission, missing ART or non-adherence, previous tuberculosis diagnosis, previous diabetes and hypertension diagnoses, family history of diabetes and hypertension, and viral load were associated with TB, HPT and T2D comorbidities among PLWH (Table 1 and 2)

### Factors associated with comorbidity of TB and Lifestyle diseases among PLWH

Table 3 summarize the baselines factors associated with TB and NCDs comorbidity among study participants. In multivariate analysis of all the various possible comorbidity combinations such ass HIV/TB, HIV/T2D, HIV/HPT, HIV/MDR TB, HIV/TB/T2D, HIV/TB/HPT, HIV/T2D/HPT and HIV/TB/T2D/HPT and baseline variable on HHIV/TB comorbidity was found significant. In multivariable model, participants who were currently taking ARV only were less likely to have HIV/TB coinfection than those taking ARV plus other medications (aOR 0.5; 95% CI 1.4 – 0.6, p = 0.0001). Further participants who were virological suppressed were less likely to have HIV/TB coinfection than those who had virological failure (uOR 0.8; 95% CI 0.6 – 0.9, p = 0.017). Participants who were previously admitted to the hospital for an ailment remained more likely to have HIV/TB coinfection than those without previous hospital admission (aOR 1.2; 95%CI 1.1 – 1.4, p = 0.049). Participants who had previous TB infection remained more likely to have HIV/TB coinfection than those without TB diagnosis (uOR 1.6; 95%CI 1.0 – 3.0, p = 0.034).

**Table 3:** Bivariable and multivariable logistic regression analysis for the covariates of HIV/TB comorbidity (n =280 patients)

| Variables | Total | n | % | HIV / TB |  |  |  |
| --- | --- | --- | --- | --- | --- | --- | --- |
|  |  |  |  | Bivariate<br>uOR (95% CI) | P - value | Multivariate<br>aOR (95% CI) | P - value |
| **Age Group** |  |  |  |  |  |  |  |
| 18 -30 | 82 | 12 | 15 | 0.8(0.2 - 2.4) | 0.651 | 0.8(0.2 - 2.5) | 0.691 |
| 31 - 40 | 121 | 27 | 22 | 0.8(0.3 - 2.6) | 0.726 | 0.8(0.3 - 2.6) | 0.778 |
| 41 - 50 | 53 | 11 | 21 | 0.8(0.3 - 2.7) | 0.714 | 0.8(0.3 - 2.7) | 0.743 |
| ≥51 | 24 | 7 | 29 | Referent | Referent | Referent | Referent |
| **Gender** |  |  |  |  |  |  |  |
| Male | 146 | 29 | 20 | 0.9(0.8 - 1.2) | 0.892 | 0.9(0.8 - 1.2) | 0.879 |
| Female | 134 | 29 | 22 | Referent | Referent | Referent | Referent |
| **Education level** |  |  |  |  |  |  |  |
| Primary | 47 | 8 | 17 | 1.1(0.7 - 1.4) | 0.978 | 1.0(0.7 - 1.4) | 0.966 |
| Secondary | 88 | 23 | 26 | 1.1(0.8 - 1.4) | 0.558 | 1.1(0.8 - 1.4) | 0.587 |
| Tertiary | 54 | 12 | 22 | 1.0(0.8 - 1.4) | 0.759 | 1.0(0.8 - 1.4) | 0.813 |
| Non-Formal | 91 | 15 | 16 | Referent | Referent | Referent | Referent |
| **Duration living with HIV (Years)** |  |  |  |  |  |  |  |
| 1 to 5 | 94 | 22 | 23.4 | 0.9(0.6 - 1.5) | 0.848 | 1.2(0.7 - 1.5) | 0.898 |
| 6 to 10 | 165 | 30 | 18.2 | 0.9(0.6 - 1.4) | 0.682 | 1.2(0.7 - 1.5) | 0.782 |
| > 11 | 21 | 6 | 28.6 | Referent | Referent | Referent | Referent |
| **ARV regimen** |  |  |  |  |  |  |  |
| ARV only | 222 | 2 | 0.9 | **0.5(0.4 - 0.6)** | **0.0001** | **0.5(0.4 - 0.6)** | **0.0001** |
| ARV plus other medications | 58 | 56 | 97 | Referent | Referent | Referent | Referent |
| **Changed ARV** |  |  |  |  |  |  |  |
| Yes | 196 | 46 | 23.5 | 1.0(0.8 - 1.3) | 0.87 | 1.3(0.6 - 1.7) | 0.988 |
| No | 84 | 12 | 14.3 | Referent | Referent | Referent | Referent |
| **Hospital Admission** |  |  |  |  |  |  |  |
| Yes | 28 | 12 | 42.9 | **1.2(1.1 - 1.7)** | **0.004** | **1.2(1.1 - 1.4)** | **0.049** |
| No | 252 | 46 | 18.3 | Referent | Referent | Referent | Referent |
| **Missed taking current ART** |  |  |  |  |  |  |  |
| Yes | 104 | 23 | 22.1 | 1.1(0.8 - 1.3) | 0.87 | 1-1(0.8 - 1.3) | 0.857 |
| No | 176 | 35 | 19.9 | Referent | Referent | Referent | Referent |
| **Duration of cough experienced** |  |  |  |  |  |  |  |
| <3 Weeks | 143 | 33 | 23.1 | 1.1(0.6 - 1.9) | 0.631 | 1.1(0.7 - 1.9) | 0.629 |
| 4-10 Weeks | 124 | 24 | 19.4 | 1.1(0.7 - 1.9) | 0.713 | 1.1(0.6 - 1.9) | 0.714 |
| >11 Weeks | 13 | 1 | 7.7 | Referent | Referent | Referent | Referent |
| **Previous tuberculosis diagnosis** |  |  |  |  |  |  |  |
| Yes | 6 | 6 | 100 | **1.6(1.0 - 2.9)** | **0.049** | **1.6(1.1 - 3.0)** | **0.034** |
| No | 274 | 52 | 19 | Referent | Referent | Referent | Referent |
| **Previous diabetes diagnosis** |  |  |  |  |  |  |  |
| Yes | 22 | 4 | 18.2 | 0.9(0.7 - 1.5) | 0.91 | 0.9(0.6 - 1.5) | 0.876 |
| No | 258 | 52 | 20.2 | Referent | Referent | Referent | Referent |
| **Body mass index (Kg/m<sup>2</sup>)** |  |  |  |  |  |  |  |
| <18.5 (Underweight) | 4 | 0 | 0 | ND | ND | ND | ND |
| 18.5 to 24.9 (Normal weight) | 132 | 25 | 18.9 | 0.7(0.3 - 1.0) | 0.461 | 0.9(0.7 - 1.2) | 0.507 |
| 25 to 29.9 (Overweight) | 92 | 16 | 17.4 | 0.9(0.7 - 1.2) | 0.438 | 0.9(0.7 - 1.2) | 0.526 |
| ≥30 (Obese) | 52 | 17 | 32.7 | Referent | Referent | Referent | Referent |
| **Baseline CD4+ (cell/μL)** |  |  |  |  |  |  |  |
| 1-500 | 186 | 38 | 20.4 | 0.9(0.8 - 1.2) | 0.951 | 1.0(0.8 - 1.2) | 0.899 |
| >501 | 94 | 20 | 21.3 | Referent | Referent | Referent | Referent |
| **HIV viral load (Cells/mls)** |  |  |  |  |  |  |  |
| 1-1000 | 246 | 44 | 17.9 | **0.04(0.05 - 0.06)** | **0.048** | **0.8(0.6 - 0.9)** | **0.017** |
| >1001 | 34 | 14 | 41.2 | Referent | Referent | Referent | Referent |
| **Baseline ALT (U/L)** |  |  |  |  |  |  |  |
| <56 | 254 | 54 | 21.3 | 1.1(0.7 - 1.5) | 0.795 | 1.1(0.7 - 1.5) | 0.799 |
| ≥56 | 26 | 4 | 15.4 | Referent | Referent | Referent | Referent |
| **Baseline AST (U/L)** |  |  |  |  |  |  |  |
| <40 | 228 | 49 | 21.5 | 1.1(0.8 - 1.4) | 0.804 | 1.0(0.8 - 1.4) | 0.797 |
| ≥40 | 52 | 9 | 17.3 | Referent | Referent | Referent | Referent |
| **Baseline HB (g/dL)** |  |  |  |  |  |  |  |
| <13 | 83 | 16 | 19.3 | 0.9(0.8 - 1.2) | 0.887 | 0.9(0.8 - 1.2) | 0.915 |
| ≥13 | 197 | 42 | 21.3 | Referent | Referent | Referent | Referent |
| **Creatinine** |  |  |  |  |  |  |  |
| <0.8mg/dL | 139 | 26 | 18.7 | 0.9(0.8 - 1.2) | 0.761 | 0.9(0.8 - 1.2) | 0.764 |
| >0.8mg/dL | 141 | 32 | 22.7 | Referent | Referent | Referent | Referent |

n - number; % - percentage; uOR – Unadjusted odd ratio; aOR – adjusted odd ratio

## Discussion

The role of NCDs as a leading global public health problem in the general population has been established. NCDs impacts PLWH significantly, and therefore regulating the increasing burden of NCDs in low and middle-income countries is imperative and it involves establishing adequate systems for monitoring the same and using the data obtained to upgrade or implement control strategies. This study provides additional data on NCDs and TB among PLWH in the eastern part of Kenya, where data is currently skewed.

The comorbidity of NCDs and TB among PLWH varied widely. The prevalence of 20.7% was reported for HIV and TB comorbidity which is lower than 37.9% reported in Ethiopia [17], 30.1% in a study in South Africa [18]. The current study reported higher prevalence of TB/HIV comorbidity than those reported in India 3.6% [19]; 11% in Tanzania [20], 2.5% in Mexico and 5.6% in the Netherlands [21, 22]. There were 2.1% PLWH coinfected with MDR-TB. This study did not report MDR-TB coinfection with either T2D or HPT. Multi drug resistant tuberculosis continues to be a global public health challenge, particularly in sub-Saharan Africa, increasing the weight of other communicable and non-communicable diseases ragging the region [23, 24]. In 2021 WHO estimates the global burden of new TB cases with MDR/RR-TB at 3.6% with 18% of them treated [24]. In a meta-analysis of Salari et al., [25], the global pooled prevalence of multi drug-resistant, Isoniazid, Rifampicin, and extensively drug-resistant TB were calculated as 11.6%, 15.7%, 9.4%, and 2.5%, respectively. In 2021, WHO further listed India, the Russian Federation and Pakistan accounting for 26%, 8.5% and 7.9% respectively of global cases of drug resistant TB burden [24]. In Pakistan Ali et al., [26] reported a prevalence of 4.9% MDR-TB cases, while in Nigeria prevalence of 3.5% MDR-TB was reported [27]. In Saudi Arabia, Sambas et al., [2020] reported a prevalence of 5% MDR TB among PLWH. The prevalence of MDR-TB seems heterogenous but the burden seems more in countries categorized by WHO as such [24]. With MDR-TB reported in this region of Kenya reiterate the need to strengthen further monitoring of TB and MDR-TB within the national HIV programs.

About 15% of the study patients had HIV/T2D comorbidity which comparable to 13.7% in Vietnam and 13.5% in Ethiopia [29, 30]. Much lower HIV/T2D coinfection rates of 11.5%, 7%, 6.1% and 5.5% have been reported in Ethiopia, Sweden, South Africa and Mali respectively [31–34]. This study reported a prevalence of 11.8% for HIV/HPT coinfection which was lower than 25.5% reported in South Africa [33], 24.8% in Uganda [35], 31.7% in Nigeria [36], 36% in France [37], and 38% in the USA [38]. Generally fewer lower prevalence of hypertension and HIV coinfection have also been reported such as 8% in Cote d’Ivoire [39]. These disparities in hypertension prevalence in PLWH may reflect different levels of traditional risk factors for hypertension in these countries.

The current prevalence of 6.4% HIV/T2D/HPT coinfection was lower than 22.6%, 19.6% and 19% reported in South Africa, Zimbabwe and Ethiopia respectively [15, 13]. The prevalence of HPT and T2D among PLWH is on the raise among African population are the leading risk factors for mortality [40, 13]. There were 3.2% of the PLWH with TB/T2D coinfection reported which was slightly lower than 5.8% and 4.8% reported in Cameron and Ethiopia respectively [41, 31]. From literature, this is among the few studies to examine the TB/HIV/T2D coinfection in Africa. Studies shows that patients with T2D are 3.59 times at risk of developing TB [42]. The burden of T2D and HIV among TB cases could be due to the fact that HIV compromises the immune system and leads to the progression to active TB among those infected with latent *M. tuberculosis* [43].

There were 3.2% PLWH in this study with HIV/TB/HPT. In a previous study in Western Kenya, a higher prevalence of HIV/TB/HPT of 11.2% and 7.4% was reported in men and women respectively [44], with 12.5% reported in Tanzania [45]. A slightly lower rates of 1.6% was reported in South Africa [46]. This study further showed that 1.4% of PLWH had quadruple comorbidity of HIV/TB/T2D and HPT. From literature data is further skewed reporting quadruple coinfection of diabetes mellitus, cardiovascular disease (e.g. hypertension), respiratory diseases (e.g. TB and chronic obstructive pulmonary diseases and pneumonia) among both the general population and PLWH. In a study conducted in the UK by Lorenc et al., [47] showed that the mean number of general and HIV-associated comorbidities amongst HIV patients was 1.1 and 1.4, respectively. The study identified diabetes, hypertension and TB most common comorbidities amongst patients with HIV. These studies show significant heterogeneity in the prevalence of TB and NCDs among PLHW. These difference in these rates could partly be explained by the overall prevalence of TB and HIV which is higher in developing countries including Kenya. HIV infected persons are 15–22 times more likely to develop active TB than HIV negative persons [48]. In the individual host, the two pathogens, *M. tuberculosis* and HIV, potentiate one another, accelerating the deterioration of immunological functions [1, 49]. Advancing age, male gender, longer duration of HIV infection, low CD4 count, high viral burden, high body mass index, greater waist circumference or waist- to-hip ratio, lower socio-economic class, and certain ethnic backgrounds or culture are among factors associated with development of NCDs among PLWH [50].

In this study only HIV/TB comorbidity were found associated with various variables evaluated. The PLWH receiving only ARV medication were protected from TB coinfection compared to those taking ARVs in combination with other mediations. This is similar to previous studies showing timely access to ART minimizes immune deterioration and improves TB outcomes [51]. Starting HAART during TB treatment is complicated by overlapping toxicities, drug interactions and immune reconstitution disease, and high pill burdens may reduce adherence [52]. Delaying HAART may lead to prolonged or worsening immune suppression. Balancing these risks when deciding when to initiate HAART with early treatment initiation reduces morbidity and mortality.

Virological suppression was protective to TB/HIV coinfection, similar to results in Turkey [53] and South Africa [54]. Association of virologic failure on ART and a higher risk for TB development has been shown [55]. It may be due to the fact that HIV accelerates the loss of CD4+ T lymphocyte and promotes the progression [53].

Previous hospital admission was associated with TB coinfection among PLWH. This was similar to a study done in Congo by Shah et al., [19] which showed that TB/HIV coinfection increases the risk of negative outcomes for PLWH, including death, loss to follow-up, and lack of viral load suppression necessitating hospitalization. In India, hospital admission was either a contributor to TB coinfection especially among the PLWH or that hospital was necessitated by TB/HIV coinfection. The study showed that majority of the patients showed improvement on being discharged, while there were about 3% who died during hospitalization. A report in India have reported high 15%, deaths for TB/HIV coinfected during hospitalization [56, 57]. Hospitalization is generally as a consequence of worsening health condition. For patients hospitalized due to TB/HIV coinfection are faced with increased risk of death may be due to treatment failure, a lack of adherence to anti-TB treatment, or a late diagnosis of TB and/or HIV [58].

Previous TB infection was associated with HIV/TB coinfection. Horsburgh et al., [59] showed that the action of previous TB infection could on one hand mount substantial protection against reinfection of TB, while at the same time observing that 36–79% of the TB disease cases can be attributed to the already-infected population. Higher rates of reactivation and reinfection are common in countries with a high incidence of TB [60].

Some other studies have identified factors associated with TB/HIV coinfection that the current study did not find observe including underlying diseases, immunosuppressive agents, substance abuse, smoking, behavioural, social and environmental factors [53]. The ratio of neutrophil to lymphocyte has also been shown as a predictor of TB among PLWHA [61].

## Conclusions

This study shows that Eastern part Kenya is experiencing a syndemic of NCDs and TB including resistant strains among PLWH denoting the need to integrate the management of NCDs among HIV and TB treatment programs in Kenya.

## Data Availability

The data has been uploaded as supplementary information.

## Acknowledgements

The authors wish to acknowledge all the study patients and staff of Level 5 Meru Hospital and Center for Microbiology Research, Kenya Medical Research Institute

## Supporting information

**S1 Data**

